# Measuring Population Health Dynamics and Health Care Costs in Medicaid Managed Care Using CareMaps

**DOI:** 10.64898/2026.02.03.26345472

**Authors:** Jacob Kuriyan, Reza Mehran

## Abstract

**Importance:** Prevention-focused health policy requires analytic frameworks capable of detecting changes in population health and associated costs within policy-relevant time horizons, particularly in managed care systems where premiums reflect actuarial risk rather than realized medical expenditures.

**Objective:** To evaluate a healthstate–based analytic framework (CareMaps) for measuring population health dynamics, disease progression, and associated costs using longitudinal Medicaid managed care claims data.

**Design, Setting, and Participants:** Retrospective longitudinal analysis of deidentified Medicaid managed care claims in New Mexico from 2011 through 2014. The study included individuals aged 0 to 64 years enrolled in managed care plans.

**Exposures:** Chronic disease burden categorized into mutually exclusive, ordered healthstates based on the number of chronic conditions.

**Main Outcomes and Measures:** County- and managed care organization (MCO) level prevalence of healthstates, transition rates between healthstates, and healthstate-specific cost estimates derived from capitation premiums and medical loss ratio defined medical expenditures.

**Results:** The CareMaps framework identified specific geographic and MCO level variation in chronic disease prevalence, healthstate transition rates, and per-member spending patterns that were not fully explained by actuarial risk adjustment. Transitions from nonchronic to chronic healthstates varied markedly across counties, indicating heterogeneity in disease progression and prevention related outcomes.

**Conclusions and Relevance:** A healthstate based analytic framework applied to longitudinal Medicaid managed care data enables standardized measurement of population health dynamics and associated costs within policy relevant time horizons. Such approaches may support evaluation of preventive care performance, inform risk adjustment, and enhance public-sector oversight of managed care programs.

## Introduction

Ninety percent of the nation’s approximately $4.9 trillion in annual health care expenditures are attributable to individuals with chronic physical and mental health conditions.^1^ Because most chronic conditions are incurable, prevention and delayed disease progression represent the primary long-term strategies for improving population health and controlling health care spending. For individuals already living with chronic illness, preventive efforts focus on reducing the risk of developing additional comorbidities that require increasingly intensive and costly interventions.

Recent federal initiatives underscore the importance of shifting from episodic treatment to longitudinal chronic care management. Programs such as the Centers for Medicare & Medicaid Services (CMS) ACCESS model emphasize coordinated, prevention-oriented care delivery across clinical and community settings.^2^

Effective prevention requires the ability to measure population health at a given point in time, track changes longitudinally, and relate those changes to economic outcomes. Cost therefore represents a third essential dimension. Preventive programs must be affordable, and policymakers must be able to distinguish those that are most likely to produce measurable improvements in population health within realistic time horizons.

In managed care systems, payments are not made for discrete services but are structured to cover the risk of future medical events. Premiums—often expressed as per-member per-month (PMPM) payments—are calculated actuarially for groups of individuals defined by geography or eligibility category. Within these groups, however, medical needs vary widely, leading to substantial heterogeneity in utilization and cost. As a result, actuarial pricing frequently defaults to prior-year premiums with incremental age-based adjustments, limiting the interpretability of premiums as indicators of population health.

The year 2014 marked the first year of Medicaid expansion under the Affordable Care Act (ACA) in 34 states, including New Mexico. Eligibility thresholds increased from 100% to 138% of the federal poverty level, resulting in enrollment of more than 125 000 additional individuals in New Mexico alone.^3^ Expansion raised concerns that newly insured individuals—many with limited prior access to care—would present with substantial unmet medical needs and impose increased strain on the health care system.

The objective of this study was to evaluate a healthstate–based analytic framework capable of measuring population health dynamics and associated costs using longitudinal Medicaid managed care claims data, with the goal of supporting prevention-oriented policy evaluation in a cost-effective manner.

## Methods

### Analytic Framework

We used CareMaps, a patented chronic population analytics platform, to evaluate managed care claims data.^4^ CareMaps is designed to support population-level monitoring of care delivery by generating geographic visualizations of disease prevalence, incidence, transitions, and treatment related costs across regions and over time.

### Healthstate definition

CareMaps segments a population into mutually exclusive, ordered healthstates based on the number of chronic conditions present: 0d (no chronic conditions), 1d (one chronic condition), 2d+ (two or more chronic conditions), and cancer (highest-severity healthstate). The number of chronic conditions serves as a proxy for disease burden and clinical risk.

Chronic conditions in this study were defined as diabetes, heart disease, hypercholesterolemia, hypertension, asthma, osteoarthritis, and cancer, and were selected based on prevalence and policy relevance in the New Mexico Medicaid population. Individuals without these conditions were classified as nonchronic, even if they had less prevalent conditions not included in the model. (Although cancer is not traditionally classified as a chronic disease, its clinical and financial impact warranted classification as the highest-severity healthstate).

### Transitions and Metrics

Over time, individuals may develop additional chronic conditions and transition from lower-to higher-severity healthstates. Because chronic diseases are generally incurable in claim-based data, transitions were treated as unidirectional. Two core population metrics were calculated: (1) the distribution of individuals across healthstates (baseline population health) and (2) transition rates between healthstates (changes in population health).

### Cost Measurement

In managed care systems, premiums reflect actuarial risk rather than realized treatment costs. By stratifying enrollees into healthstates with broadly similar medical needs, CareMaps enables estimation of healthstate-specific costs using premiums as proxies. This approach enables comparison of economic patterns associated with disease progression across counties and managed care organizations.

### Study Population and Analysis

We analyzed deidentified medical, hospital, and pharmacy claims for New Mexico Medicaid managed care enrollees from 2011 through 2014, using 2010 data to establish baseline chronic disease status. Individuals aged 65 years or older, those enrolled in long-term care, and those receiving specialized non–chronic-related services were excluded. Analyses were stratified by age group (0–49 years and 50–64 years). Counties with small sample sizes were aggregated into super-counties to stabilize estimates.

## Results

Across a predominantly younger Medicaid managed care population, the distribution of enrollees by healthstate demonstrates substantial prevention opportunity, with the majority of beneficiaries residing in nonchronic or single–chronic-condition categories (Figure 1). This population structure establishes a favorable context in which upstream interventions could plausibly alter long-term disease trajectories and spending growth before multimorbidity becomes prevalent.

**Figure 1.**
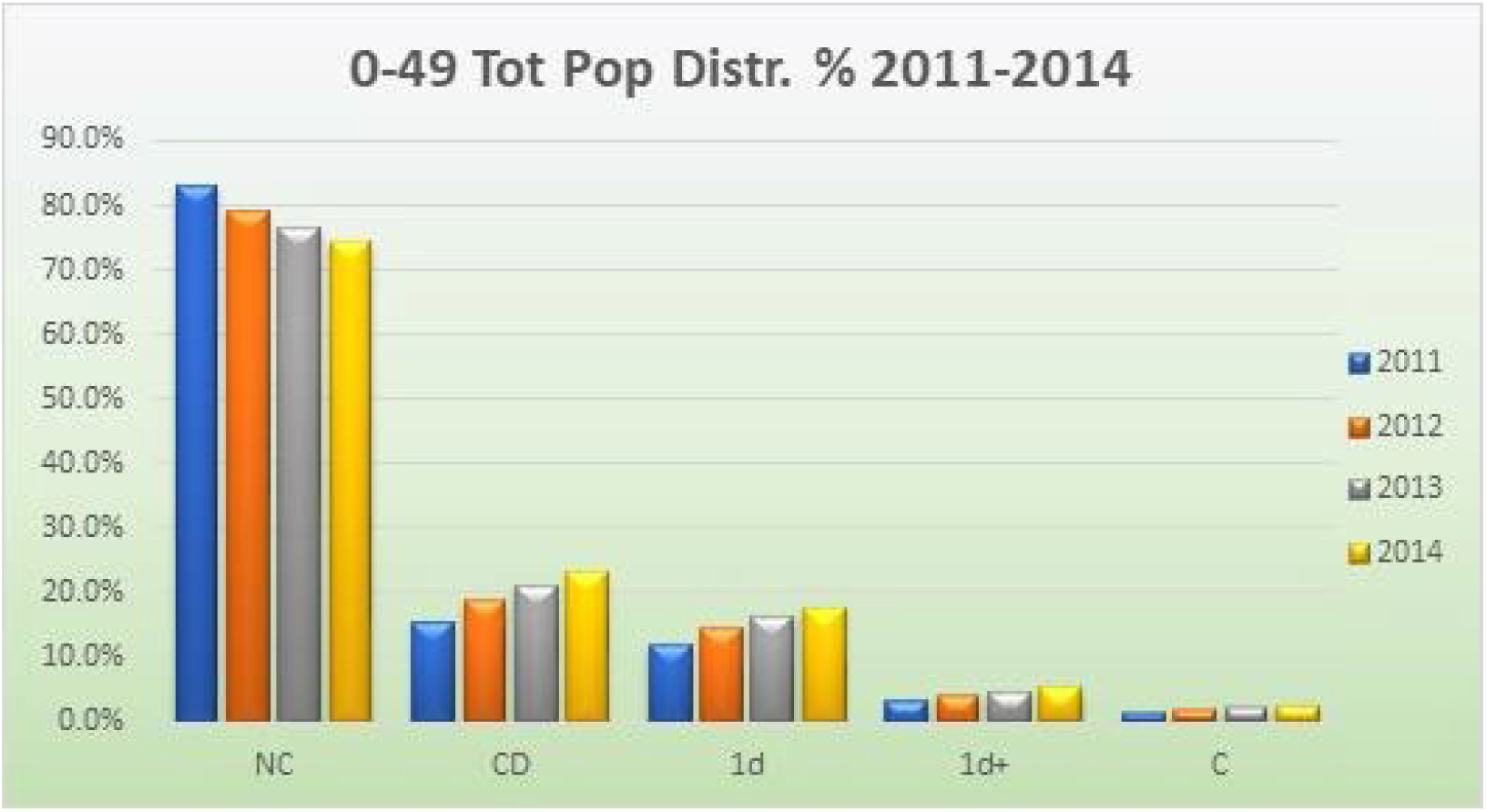
Total population distribution for the age group 0 to 49. NC is “0d” or no chronic condition; CD is for one or more chronic condition; “1d” is for one chronic condition; 1d+ for 2 or more chronic conditions; and C is Cancer

**Figure 2.**
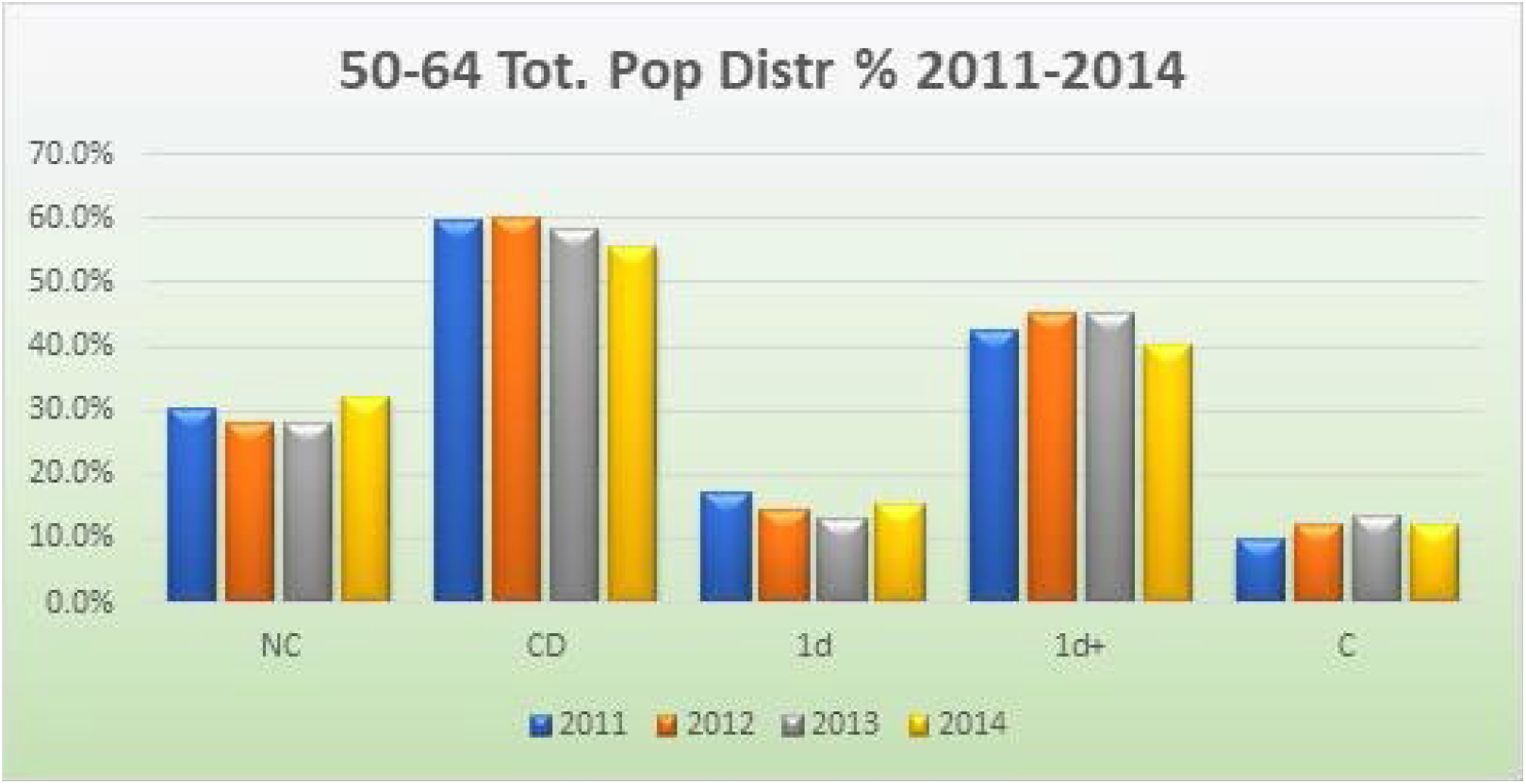
Total population distribution for the age group 50-64. NC is “0d” or no chronic condition; CD is for one or more chronic condition; “1d” is for one chronic condition; 1d+ for 2 or more chronic conditions; and C is Cancer

Despite comparable disease burden within defined healthstates, per-member per-month (PMPM) premiums vary markedly across counties and super-counties, indicating that geographic price dispersion is not fully explained by underlying clinical risk (Figure 3). This pattern persists and intensifies as disease severity increases, with premium variability expanding substantially among enrollees with multiple chronic conditions or cancer (Figure 4). The amplification of dispersion in higher-severity healthstates suggests that pricing mechanisms may be increasingly decoupled from standardized risk as clinical complexity rises.

**Figure 3.**
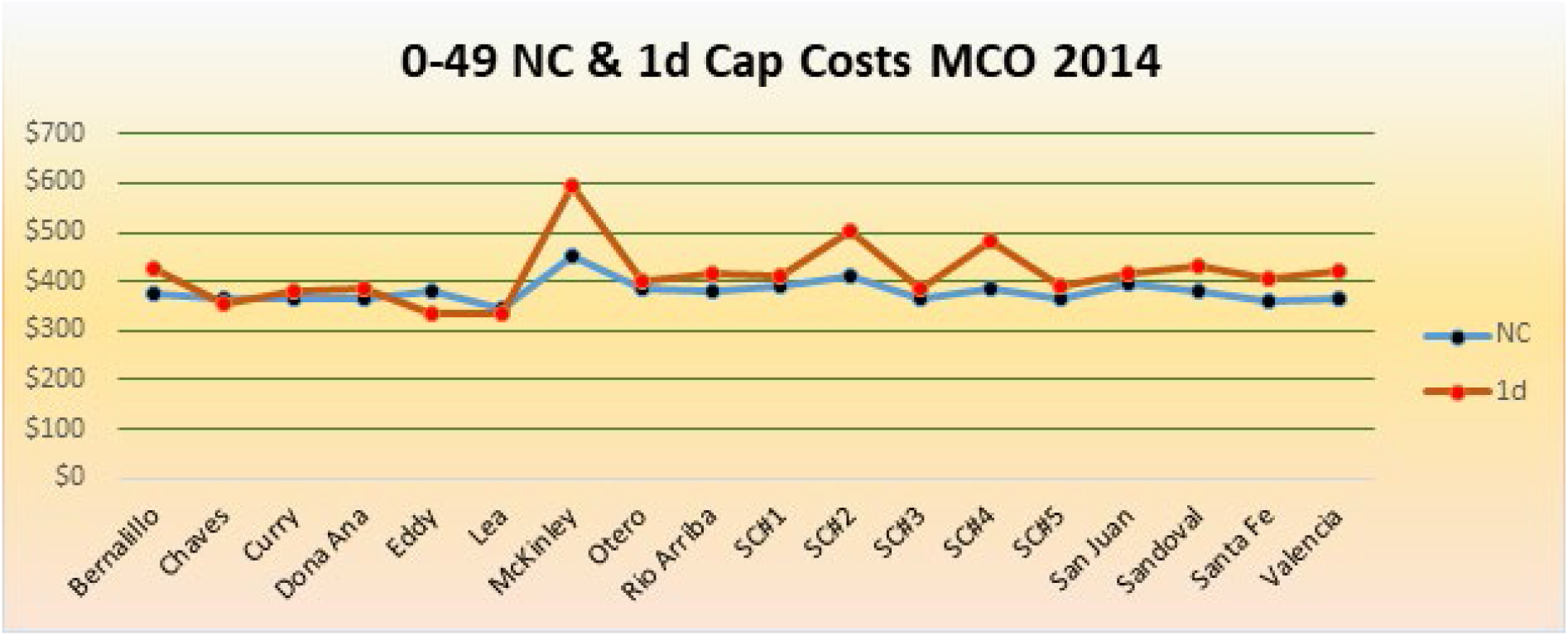
County-Level Premium Variation for Nonchronic (0d) and Single-Condition (1d) Healthstates. SC refers to “Super Counties”. MCO: Managed Care Organizations.

**Figure 4.**
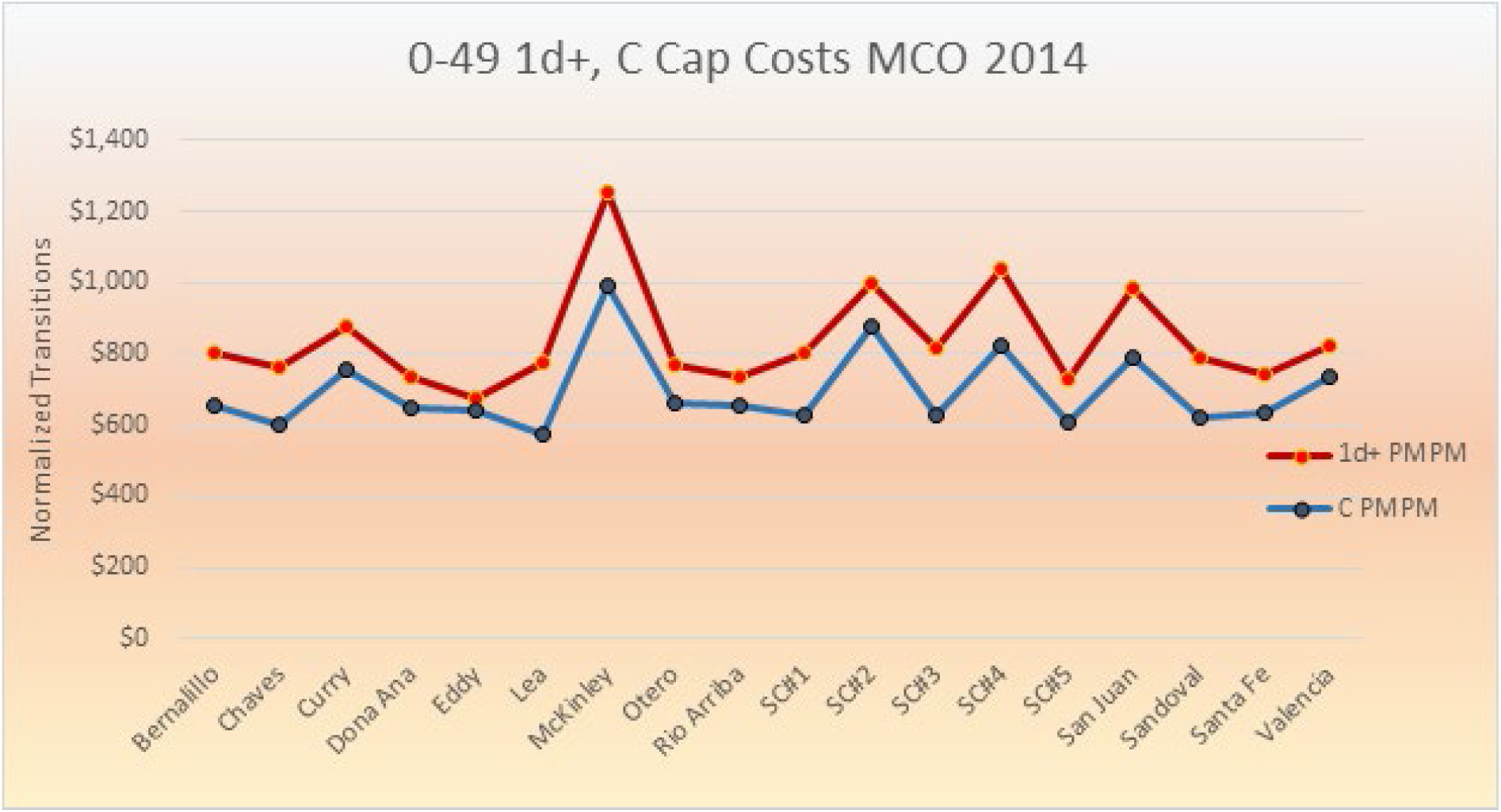
1d+ and C Premium Variations. 1d+: one heathsate or more. C: Cancer. SC: “Super Counties”, groups of smaller counties to make total enrollment statistically significant. PMPM: Payment per member. MCO: Managed Care Organizations.

Plan-level analyses further reinforce this conclusion. Within the same counties and among enrollees in identical healthstates, PMPM premiums differ meaningfully across Medicaid managed care organizations, reflecting organizational rather than population-driven variation in payment (Figure 5). Along with county-level findings, these results point to structural differences in contracting, pricing strategies, or administrative efficiency that operate independently of enrollee health status.

**Figure 5.**
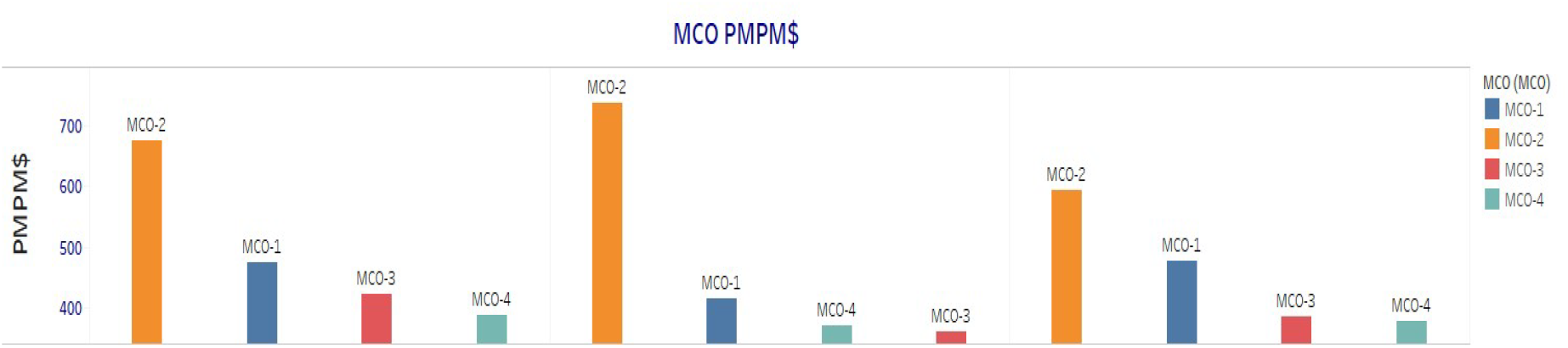
MCO premium PMPM for 3 sample counties, Bernalillo, Dona Ana and Santa Fe, for the 4 different MCOs. MCO: Managed Care Organization. PMPM: Payment per member.

Finally, comparison of aggregate premiums with incurred medical costs among older Medicaid enrollees approaching Medicare eligibility highlights systematic divergence from the 85% medical loss ratio benchmark (Figure 6). Observed deviations suggest both potential overpayment in some settings and excess medical expenditure in others, suggesting inefficiencies that may compound as beneficiaries age and transition into higher-cost healthstates.

**Figure 6.**
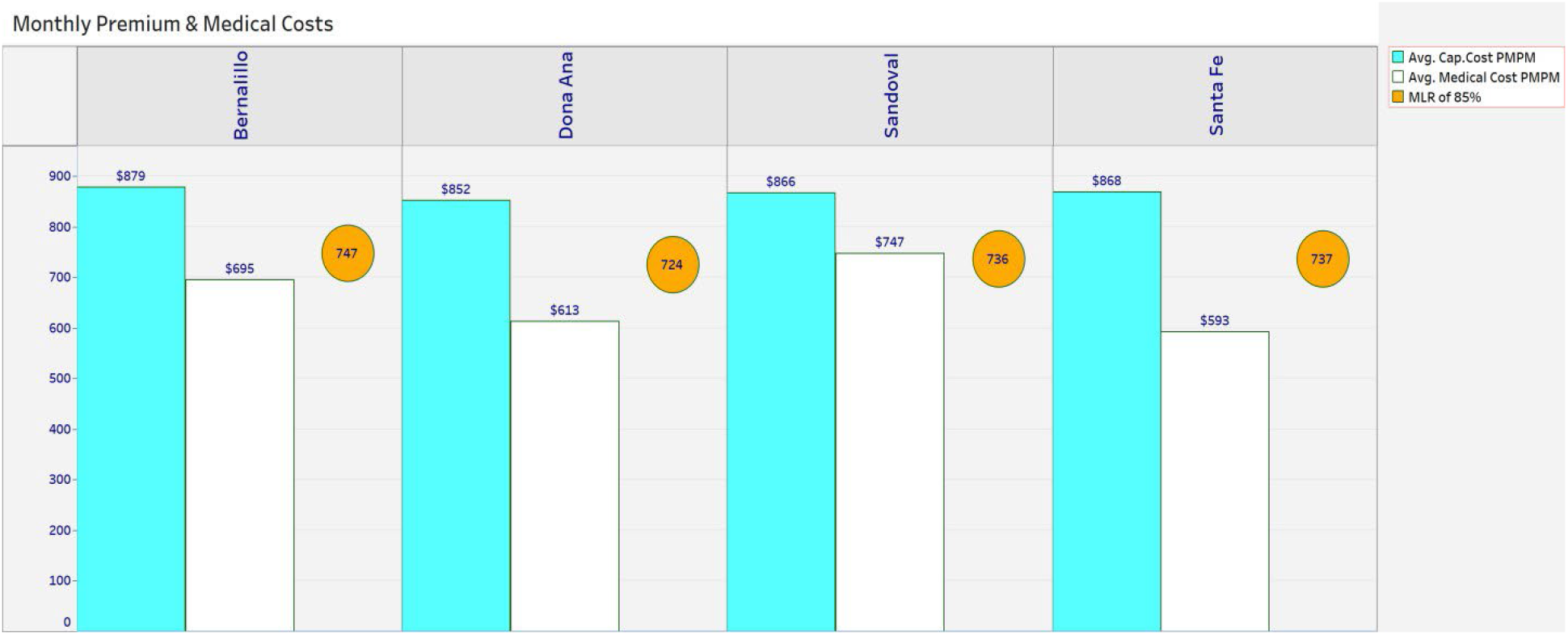
Medial Loss Ration (MLR) for 2014 and the age range 50-64, MLRs are calculated for all the four MCOs combined. The Premium and inccurred medical costs are the bar graphs (magenta and whie respectively), and the 85% value is illustrated with as a yellow circle. The data is just for 4 counties, Bernalillo, Dona Ana, Sandoval and Santa Fe counties.

## Discussion

Chronic disease prevention represents the most plausible long-term strategy for controlling health care spending, given that chronic conditions account for the majority of medical expenditures.^1^ Our findings demonstrate that a substantial proportion of Medicaid managed care enrollees reside in nonchronic or early chronic healthstates, underscoring the policy relevance of prevention in populations where disease trajectories remain modifiable. Adoption of healthy behaviors—such as physical activity, nutritious diets, and maintenance of healthy body weight— is central to prevention, yet is strongly shaped by social determinants of health and community level factors.^5^–^7^ The observed variation in premiums across counties and plans, despite comparable disease burden, suggests that current payment structures may insufficiently reflect these upstream determinants of risk.

Within Medicaid and Medicare managed care, the medical loss ratio (MLR) framework governs the allocation of premium revenue between medical and nonmedical spending. Federal regulations require that at least 85% of premiums be devoted to medical care, with deviations resulting in either required repayments or absorbed losses by managed care organizations. Our analysis demonstrates that, even within this standardized regulatory environment, meaningful divergence between premiums and incurred medical costs persists. The presence of both potential overpayment in some settings and excess medical expenditure in others indicates that premium setting and risk adjustment may remain imperfectly aligned with actual utilization patterns.

The temporary assumption of medical risk by the federal government during the initial Medicaid expansion period further highlights the importance of accurate premium calibration. Although expansion enrollees were protected from medical cost risk, nonmedical costs and profits remained excluded from federal reimbursement. This structure may have introduced or amplified incentives for pricing variation unrelated to underlying disease burden, a pattern consistent with the plan-level and geographic premium dispersion observed in this study.

Although conducted in a single State, this analysis employs a framework that is broadly applicable to other Medicaid managed care programs with comparable claims data. The CareMaps framework provides a standardized, population-level measurement infrastructure that links healthstate transitions to economically and regulatorily meaningful outcomes. By aligning premium-based estimates with MLR thresholds, the framework enhances transparency for policymakers and payers seeking to distinguish appropriate risk-based pricing from inefficiencies.

These findings suggest that improved alignment between prevention investment, premium setting, and regulatory oversight could yield durable gains in cost containment and equity. The coexistence of overpayment and excess medical spending within the same managed care environment represents not a contradiction, but an opportunity—one that can be addressed through data-driven recalibration of risk, pricing, and prevention strategy.

## Limitations

This analysis has several limitations. First, it is based on Medicaid managed care claims from a single State and a restricted time period (2010–2014), reflecting the scope of data made available by the State data custodian; more recent claims were not accessible for this analysis. Although this may limit assessment of temporal trends beyond the study period, the analytic framework itself is not time dependent and is applicable to more recent data when available. Second, chronic disease status was defined using a selected set of conditions observable in claims, potentially misclassifying some individuals as nonchronic and biasing results toward underestimation of disease burden. Third, capitation premiums were used as proxies for healthstate-specific costs, reflecting actuarial pricing rather than realized expenditures but representing the policy-relevant economic signal in managed care. Finally, claims data do not capture social or behavioral determinants, limiting causal inference.

## Conclusion

A healthstate–based analytic framework applied to longitudinal Medicaid managed care data enables standardized measurement of population health dynamics and associated costs. Premium variation in Medicaid managed care is shaped not only by disease burden but also by geography, plan-specific factors, and age-related transitions in care. Together, these effects identify a critical policy window in younger populations, where prevention-oriented strategies and improved alignment between premiums, risk, and medical spending could yield durable gains in both equity and cost containment.

## Data Availability

All data produced in the present study are available upon reasonable request to the authors.

## Tables

**Table 1:**
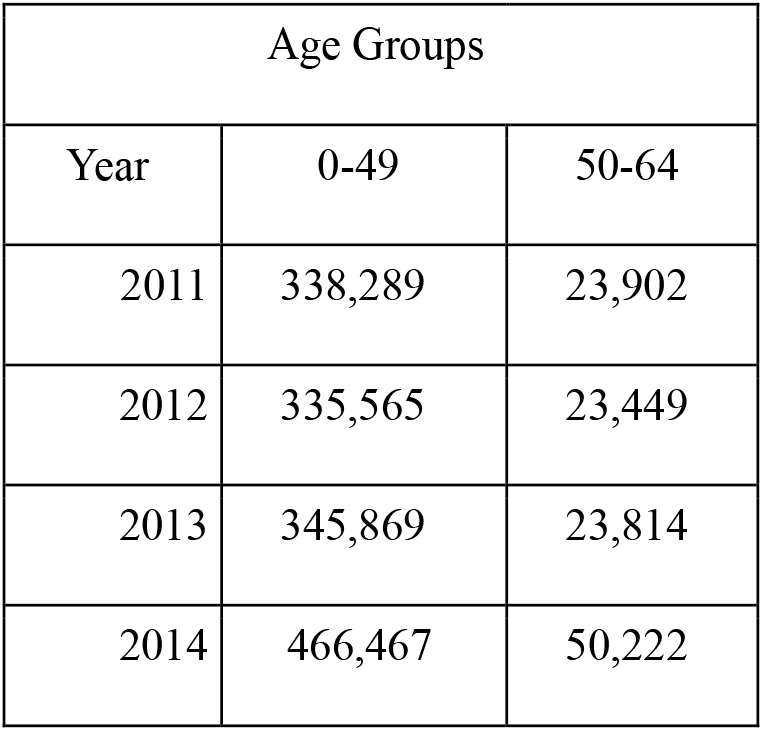
Cohort distribution in managed care enrollments.

| Age Groups |  |  |
| --- | --- | --- |
| Year | 0-49 | 50-64 |
| 2011 | 338,289 | 23,902 |
| 2012 | 335,565 | 23,449 |
| 2013 | 345,869 | 23,814 |
| 2014 | 466,467 | 50,222 |

## Figure Legends

**Figure 1. Distribution of Healthstates Among Medicaid Managed Care Enrollees Aged 0–49 Years**

Distribution of Medicaid managed care enrollees aged 0–49 years across healthstates, defined by the number of chronic conditions: nonchronic (0d), single chronic condition (1d), multiple chronic conditions (1d+), and cancer (C). This distribution provides baseline context for prevention potential and downstream cost analyses in a younger Medicaid population.

**Figure 2. Distribution of Healthstates Among Medicaid Managed Care Enrollees Aged 50– 64 Years**

Distribution of Medicaid managed care enrollees aged 50–64 years across healthstates. Compared with younger enrollees, higher-severity healthstates are more prevalent, reflecting increased disease burden and greater expected medical expenditures in this age group.

**Figure 3. County-Level Premium Variation for Nonchronic (0d) and Single-Condition (1d) Healthstates**

County-level variation in per-member per-month (PMPM) premiums for enrollees in the nonchronic (0d) and single-condition (1d) healthstates. Premiums are shown for counties and aggregated super-counties (SC), defined as groups of adjacent counties combined to ensure statistical stability. Variation is observed despite comparable disease burden within healthstates.

**Figure 4. County-Level Premium Variation for Higher-Severity Healthstates (1d+ and Cancer)**

County-level PMPM premium variation for enrollees in higher-severity healthstates (multiple chronic conditions [1d+] and cancer). Premium dispersion increases with disease burden, illustrating amplification of cost variation among populations with more complex clinical needs.

**Figure 5. Managed Care Organization–Level PMPM Premiums for Enrollees in the 1d Healthstate**

Comparison of PMPM premiums for enrollees in the single-condition (1d) healthstate across four Medicaid managed care organizations (MCOs) within selected counties (Bernalillo, Doña Ana, and Santa Fe) in 2014. Differences reflect plan-level variation among MCOs serving similar populations.

**Figure 6. Medical Loss Ratio Analysis for Medicaid Managed Care Enrollees Aged 50–64 Years**

Comparison of aggregate premiums and incurred medical costs for Medicaid managed care enrollees aged 50–64 years in 2014. Bars represent premiums and medical expenditures; the reference marker denotes the 85% medical loss ratio (MLR) threshold. Results illustrate alignment between expected and realized medical spending and identify potential overpayment or excess medical expenditures.

## Supplemental Figure Legends

**(eAppendix) eFigure 1. County-Level Prevalence of Hypertension Among Enrollees Aged 50–64 Years**

Geographic distribution of hypertension prevalence among Medicaid managed care enrollees aged 50–64 years in 2013. Counties are color-coded by Z score, representing deviation from the statewide mean prevalence.

**eFigure 2. County-Level Prevalence of Diabetes Among Enrollees Aged 0–49 Years**

Geographic distribution of diabetes prevalence among Medicaid managed care enrollees aged 0– 49 years in 2012. Counties are color-coded by Z score relative to the statewide mean.

**eFigure 3. Transitions From Nonchronic Healthstate to Diabetes Among Enrollees Aged 0– 49 Years**

County-level transition rates from the nonchronic (0d) healthstate to diabetes among Medicaid managed care enrollees aged 0–49 years in 2012, normalized to a population of 10 000 enrollees.

**eFigure 4. Transitions From Nonchronic Healthstate to Heart Disease Among Enrollees Aged 50–64 Years**

County-level transition rates from the nonchronic (0d) healthstate to heart disease among Medicaid managed care enrollees aged 50–64 years in 2013, normalized to a population of 10 000 enrollees.

**eFigure 5. Managed Care Organization–Level Distribution of the 1d Healthstate**

Distribution of enrollees in the single-condition (1d) healthstate across four Medicaid managed care organizations within selected counties (Bernalillo, Doña Ana, and Santa Fe) in 2014.

**eFigure 6. Managed Care Organization–Level Transition Rates From Nonchronic to 1d Healthstate**

Transition rates from the nonchronic (0d) healthstate to the single-condition (1d) healthstate across four Medicaid managed care organizations within selected counties in 2014, normalized to a population of 10 000 enrollees.

## References

1. Centers for Disease Control and Prevention. Fast facts: health and economic costs of chronic conditions. Accessed Month Day, Year.

2. Centers for Medicare & Medicaid Services. Unlocking a new era of chronic care: overview of the CMS ACCESS model. January 2026.

3. Medicaid Expansion Under the ACA: A State-by-State Guide. GovFacts.

4. Kuriyan J. Chronic population–based cost model to compare effectiveness of preventive care programs. US Patent 8,676,598. March 2014.

5. World Health Organization. Preventing chronic diseases: a vital investment. 2006.

6. World Health Organization. Global mapping report on multisectoral actions to strengthen the prevention and control of noncommunicable diseases and mental health conditions. 2023.

7. World Health Organization. Noncommunicable diseases progress monitor 2025. 2025.

