## Supplemental figures for "Measuring Population Health Dynamics and Health Care Costs in Medicaid Managed Care Using CareMaps"

**List of eFigures:**

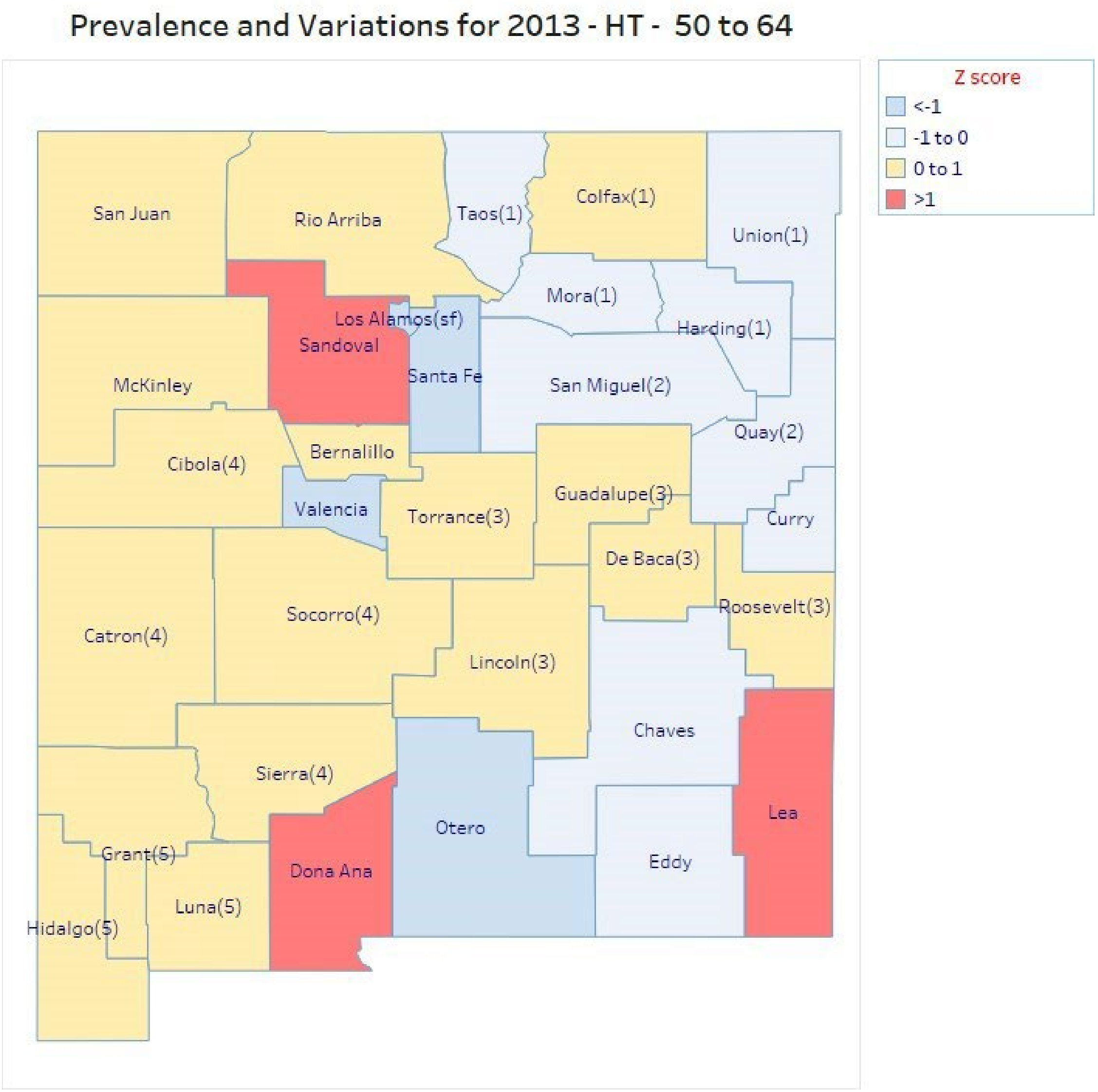


eFigure 1. Geographic distribution of hypertension (HT) prevalence among Medicaid managed care enrollees aged 50–64 years in 2013. Counties are color-coded by Z score, representing deviation from the statewide mean prevalence.

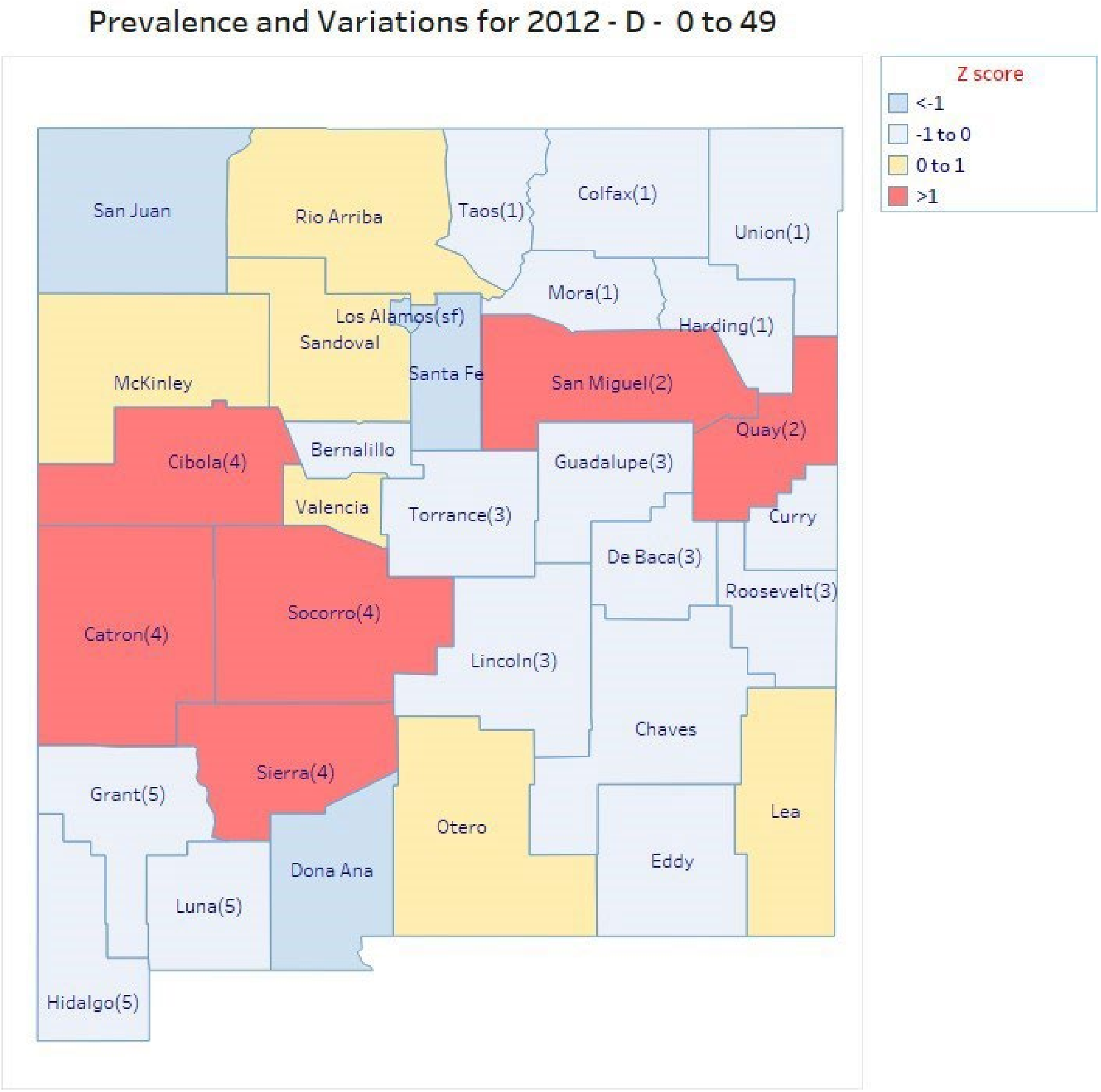


eFigure 2. Geographic distribution of diabetes (D) prevalence among Medicaid managed care enrollees aged 0–49 years in 2012. Counties are color-coded by Z score relative to the statewide mean.

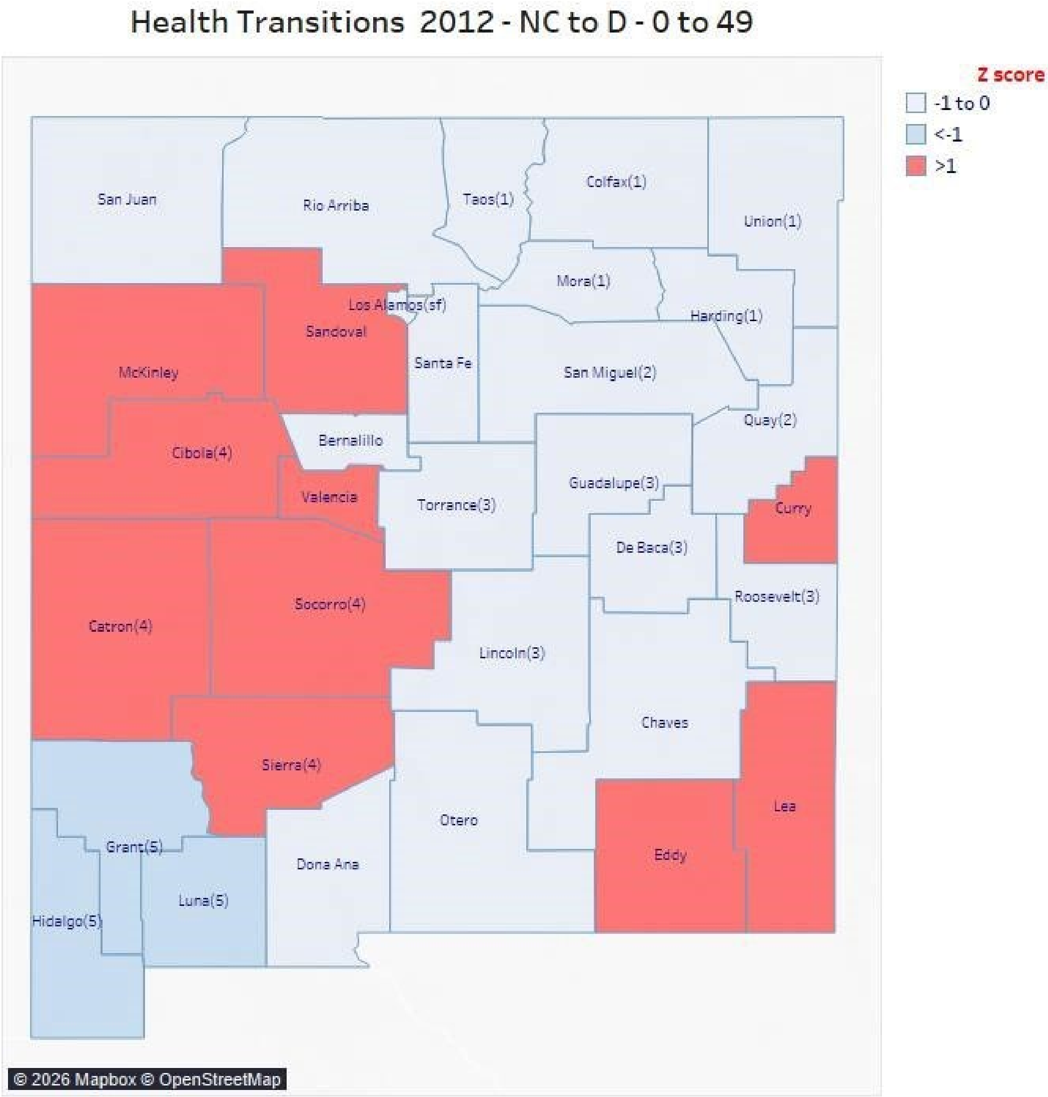


eFigure 3. County-level transition rates from the nonchronic (NC) healthstate to diabetes (D) among Medicaid managed care enrollees aged 0–49 years in 2012, normalized to a population of 10 000 enrollees.

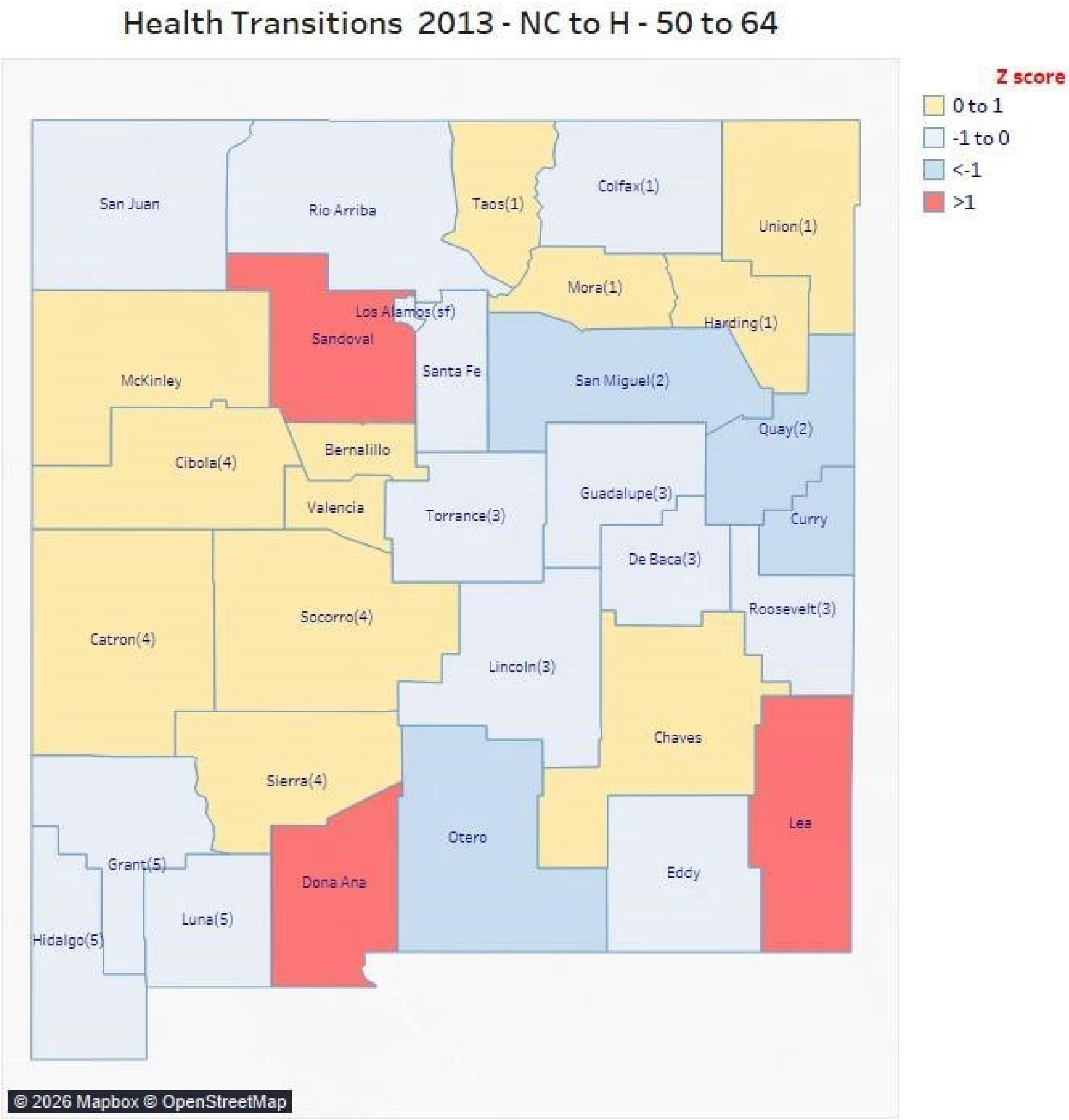


eFigure 4. County-level transition rates from the nonchronic (NC) healthstate to heart disease (H), among Medicaid managed care enrollees aged 50–64 years in 2013, normalized to a population of 10 000 enrollees.

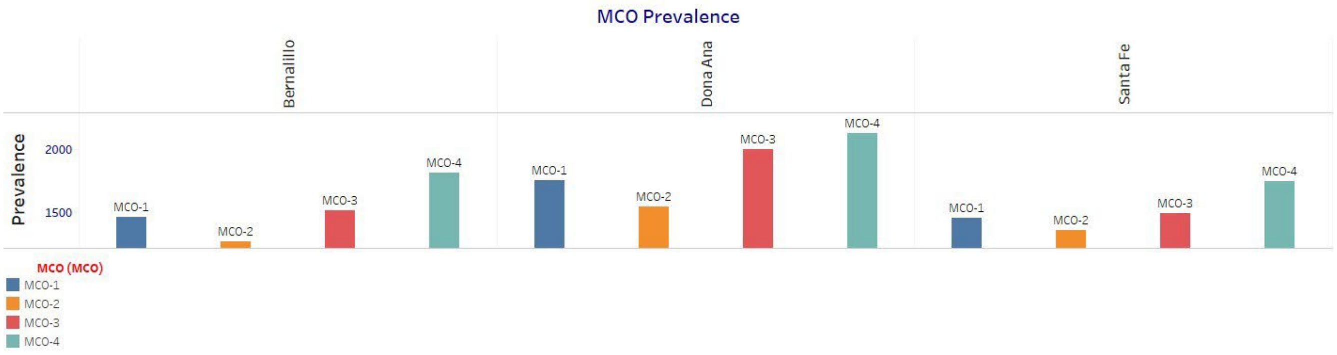


eFigure 5. Distribution of enrollees in the single-condition (1d) healthstate across four Medicaid managed care organizations within selected counties (Bernalillo, Doña Ana, and Santa Fe) in 2014.

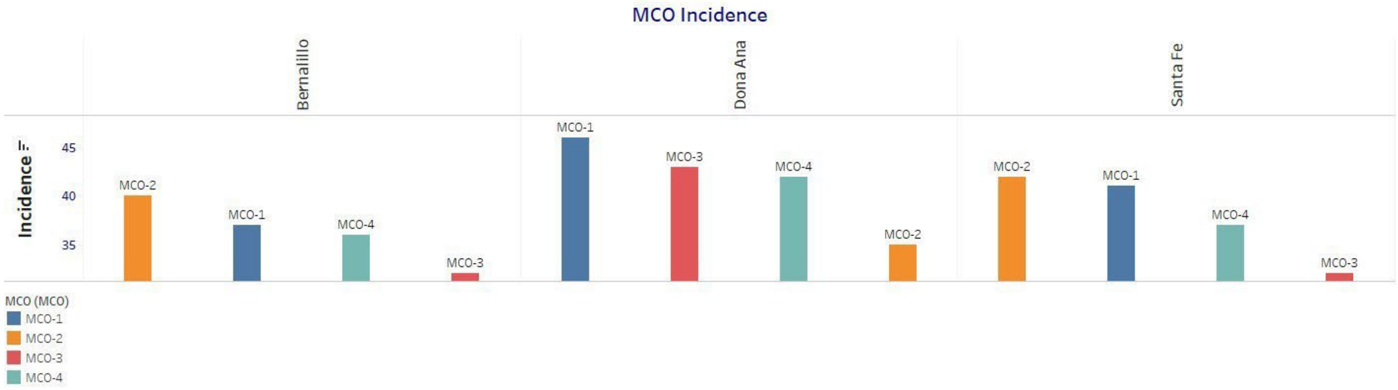


eFigure 6. Transition rates from the nonchronic (NC) healthstate to the single-condition (1d) healthstate across four Medicaid managed care organizations within selected counties in 2014, normalized to a population of 10 000 enrollees.
